## Supplementary materials for "Omega-3 supplements in the prevention and treatment of youth depression and anxiety: A scoping review"

**Supplementary Table 1. Search terms for academic databases: Search date 4^th^ August 2021**

| **Database** | **Search Terms** |
| --- | --- |
| Cochrane  CENTRAL | (“omega-3” OR “n-3” OR “ω-3” OR "polyunsaturated fatty acid*" OR "unsaturated fatty acid*" OR "fatty acid" OR "fatty acids" OR "polyunsaturated fat" OR "polyunsaturated fats" OR "polyunsaturated fatty” OR PUFA* OR "eicosapentaenoic acid" OR "docosahexaenoic acid" OR EPA OR DHA or “fish oil” or “cod liver oil”):ti,ab,kw.  **AND**  (child* or adolesc* or pediatri* or paediatri* or youth or "young person" or "young persons" or "young people" or "young peoples" or "young adult" or "young adulthood" or "young adults" or teen* or student* or high?school or undergrad* or college or university or pubescent):ti,ab,kw  **AND**  (depress* or dysthymi* or "affective disorder*" or "mood disorder*" or "anxiety disorder*" or anxi* or generali?ed anxi* or GAD or "mental health" or phobia* or panic or stress):ti,ab,kw. |
| EmBASE | omega-3 or n-3 or ω-3 or "polyunsaturated fatty acid*" or "unsaturated fatty acid*" or ("fatty acid" OR "fatty acids") or ("polyunsaturated fat" OR "polyunsaturated fats" OR "polyunsaturated fatty") or PUFA* or "eicosapentaenoic acid" or "docosahexaenoic acid" or EPA or DHA or fish oil* or cod liver oil).ti,ab.  **AND**  (child* or adolesc* or pediatri* or paediatri* or youth or ("young person" OR "young persons") or ("young people" OR "young peoples") or ("young adult" OR "young adulthood" OR "young adults") or teen* or student* or high?school or undergrad* or college or university or pubescent).ti,ab.  **AND**  (depress* or dysthymi* or "affective disorder*" or "mood disorder*" or "anxiety disorder*" or anxi* or generali?ed anxi* or GAD or "mental health" or phobia* or panic or stress).ti,ab. |
| PsycINFO | (ti(omega-3 or n-3 or ω-3 or "polyunsaturated fatty acid*" or "unsaturated fatty acid*" or ("fatty acid" OR "fatty acids") or ("polyunsaturated fat" OR "polyunsaturated fats" OR "polyunsaturated fatty") or PUFA* or "eicosapentaenoic acid" or "docosahexaenoic acid" or EPA or DHA or fish oil* or cod liver oil) OR ab(omega-3 or n-3 or ω-3 or "polyunsaturated fatty acid*" or "unsaturated fatty acid*" or ("fatty acid" OR "fatty acids") or ("polyunsaturated fat" OR "polyunsaturated fats" OR "polyunsaturated fatty") or PUFA* or "eicosapentaenoic acid" or "docosahexaenoic acid" or EPA or DHA or fish oil* or cod liver oil))  **AND**  (ti(child* or adolesc* or pediatri* or paediatri* or youth or ("young person" OR "young persons") or ("young people" OR "young peoples") or ("young adult" OR "young adulthood" OR "young adults") or teen* or student* or high?school or undergrad* or college or university or pubescent) OR ab(child* or adolesc* or pediatri* or paediatri* or youth or ("young person" OR "young persons") or ("young people" OR "young peoples") or ("young adult" OR "young adulthood" OR "young adults") or teen* or student* or high?school or undergrad* or college or university or pubescent))  **AND**  (ti(depress* or dysthymi* or ("affective disorder" OR "affective disorders") or ("mood disorder" OR "mood disorders") or ("anxiety disorder" OR "anxiety disorders") or anxi* or “generali?ed anxi*” or GAD or “mental health” or phobia* or panic or stress) OR ab(depress* or dysthymi* or ("affective disorder" OR "affective disorders") or ("mood disorder" OR "mood disorders") or ("anxiety disorder" OR "anxiety disorders") or anxi* or “generali?ed anxi*” or GAD or “mental health” or phobia* or panic or stress)) |
| PubMed | "omega-3"[Title/Abstract] OR "n-3"[Title/Abstract] OR "omega-3"[Title/Abstract] OR "polyunsaturated fatty acid*"[Title/Abstract] OR "unsaturated fatty acid*"[Title/Abstract] OR "fatty acid"[Title/Abstract] OR "fatty acids"[Title/Abstract] OR "polyunsaturated fat"[Title/Abstract] OR "polyunsaturated fats"[Title/Abstract] OR "polyunsaturated fatty"[Title/Abstract] OR "pufa*"[Title/Abstract] OR "eicosapentaenoic acid"[Title/Abstract] OR "docosahexaenoic acid"[Title/Abstract] OR "EPA"[Title/Abstract] OR "DHA"[Title/Abstract] OR "fish oil*"[Title/Abstract] OR "cod liver oil"[Title/Abstract]  **AND**  "child*"[Title/Abstract] OR "adolesc*"[Title/Abstract] OR "pediatri*"[Title/Abstract] OR "paediatri*"[Title/Abstract] OR youth[Title/Abstract] OR "young person"[Title/Abstract] OR "young persons"[Title/Abstract] OR "young people"[Title/Abstract] OR "young peoples"[Title/Abstract] OR "young adult"[Title/Abstract] OR "young adults"[Title/Abstract] OR "young adulthood"[Title/Abstract] OR teen*[Title/Abstract] OR student*[Title/Abstract] OR high?school[Title/Abstract] OR undergrad*[Title/Abstract] OR college[Title/Abstract] OR university[Title/Abstract] OR pubescent[Title/Abstract]  **AND**  "depress*"[Title/Abstract] OR "dysthymi*"[Title/Abstract] OR "affective disorder"[Title/Abstract] OR "affective disorders"[Title/Abstract] OR "mood disorder"[Title/Abstract] OR "mood disorders"[Title/Abstract] OR "anxiety disorder"[Title/Abstract] OR "anxiety disorders"[Title/Abstract] OR "anxi*"[Title/Abstract] OR "generalised anxi*"[Title/Abstract] OR "generalized anxi*"[Title/Abstract] OR GAD[Title/Abstract] OR "mental health"[Title/Abstract] OR "phobia*"[Title/Abstract] OR "panic"[Title/Abstract] OR "stress"[Title/Abstract] |

**Supplementary Table 2. Grey literature search terms and databases**

| **Database** | **Date searched** | **Search terms** |
| --- | --- | --- |
| Substance Abuse & Mental Services Administration | 4.8.21 | omega-3, n-3, ω-3, fatty acid, polyunsaturated fat, fish oil, cod liver oil |
| World Health Organization Regional Office for Europe - Health Evidence Network | 4.8.21 | omega-3, n-3, ω-3, fatty acid, polyunsaturated fat, fish oil, cod liver oil |
| Canadian Agency for Drugs and Technologies in Health | 4.8.21 | omega-3, “ω-3” “polyunsaturated fatty acid” “unsaturated fatty acid" fatty acid” “polyunsaturated fat” "fish oil" |
| International Health Technology Assessment database | 10.8.21 | (omega-3 or n−3 “ω-3” or “polyunsaturated fatty acid*” or “unsaturated fatty acid*” or “fatty acid” or “polyunsaturated fat*” or PUFA* or “eicosapentaenoic acid” or “docosahexaenoic acid” or EPA or DHA or “fish oil*” or “cod liver oil” or “cod-liver oil”) AND (child* or adolesc* or pediatri* or paediatri* or youth or “young people” or “young adult” or teen* or student* or “high school” or “high-school” or undergraduate or college or university or pubescent)  AND (depress* or dysthymi* or “affective disorder*” or “mood disorder*” or “anxiety disorder” or anxiety or anxious or “mental health” or phobia* or panic or stress) |
| National Institute for Health and Care Excellence | 10.8.21 | (omega 3 or "fatty acid" or "fish oil") AND (mental health or anxiety or depression) AND ("young people" or youth or child or adolescent) |
| National Institute for Health Research Innovation Observatory | 10.8.21 | omega-3, n-3, ω-3, fatty acid, polyunsaturated fat, fish oil, cod liver oil |
| Clinical Trials Registry - India | 16.8.21 | omega-3 |
| Open trials (https://explorer.opentrials.net/) | 16.8.21 | (Fish oil OR omega-3 OR fatty acid OR polyunsaturated fat) AND (depression OR mental health OR anxiety) |
| Trip Medical Database | 17.8.21 | Omega-3, fish oil, fatty acid, depression, anxiety, mental health, adolescent, child, youth |
| Google Advanced | 17.8.21 | (omega-3 OR “ω-3” OR "fatty acid" OR "polyunsaturated fat" OR PUFA OR "cod liver oil" OR "fish oil") AND (child OR adolescent OR youth OR "young people" OR "young adult" OR teen OR student) AND (depression OR dysthymia OR anxiety OR mood OR "mental health") |

**Supplementary Table 3. Cochrane Risk of Bias ratings for randomised controlled trials (n=13)**

| Studies | Item 1:  Risk of bias arising from the randomisation process | Item 2:  Risk of bias arising from the effect of assignment to intervention | Item 3:  Risk of bias due to missing outcome data | Item 4:  Risk of bias in measurement of outcome | Item 5:  Risk of bias in selection of the reported result |
| --- | --- | --- | --- | --- | --- |
| Amminger 2010 | Low | Low | Low | Some concerns | Some concerns |
| Amminger 2013 | Low | Low | Low | Low | Some concerns |
| Gabbay 2019 | Some concerns | Low | Some concerns | Low | Low |
| Giles 2015 | Some concerns | Some concerns | Low | Some concerns | Some concerns |
| Ginty 2015 | Low | Low | Low | Low | Some concerns |
| Jamilian 2018 | Low | Some concerns | Low | Low | Some concerns |
| Kiecolt-Glaser 2011 | Low | Low | Low | Low | Low |
| Manos 2018 | Some concerns | Low | Low | Low | Some concerns |
| McGorry 2017 | Low | Low | Low | Low | Low |
| McNamara 2020 | Some concerns | Low | Some concerns | Low | Some concerns |
| Robinson 2019 | Some concerns | Low | Low | Low | Some concerns |
| Trebatická 2020 | Low | Low | Low | Low | Some concerns |
| Van der Wurff 2020 | Some concerns | Low | Low | Low | Low |

**Supplementary Table 4. Ratings of grey literature according to comprehensiveness, accuracy of information, and reference to peer-reviewed literature (n=12)**

| **Sources** | **Comprehensiveness** | **Accuracy of information** | **Reference to peer-reviewed literature** |
| --- | --- | --- | --- |
| Amen Clinics, 2020 | Excellent | Moderate | Excellent |
| British Diet Association, 2020 | Poor | Moderate | Poor |
| Contemporary Paediatrics, 2005 | Poor | Moderate | Moderate |
| Headspace, 2019 | Poor | Moderate | Poor |
| Healthline, 2019 | Moderate | Moderate | Excellent |
| Hey Sigmund, 2017 | Poor | Poor | Poor |
| Kaiser Permanente, 2021 | Excellent | Excellent | Excellent |
| Newport Academy, 2017 | Moderate | Moderate | Moderate |
| Nutri Advanced, 2021 | Excellent | Moderate | Excellent |
| Pediatric Partners, no date | Poor | Moderate | Excellent |
| UNICEF, 2015 | Poor | Poor | Excellent |
| Vital Choice, 2014 | Excellent | Excellent | Excellent |

**Supplementary Table 5. Characteristics of included non-randomised controlled trials (n=4)**

| **Study ID** | **Study type** | **Country** | **Sample size, n** | **Female  participants**  **(%)** | **Demographic** | **Diagnostic criteria** | **Age, mean (SD)** | **Conditions** | **Outcome measure** |
| --- | --- | --- | --- | --- | --- | --- | --- | --- | --- |
| **Amminger, 2015 (34)** | Pilot study | Australia | 68 | 64.7 | Adolescents experiencing moderate to severe depressive symptoms | DSM-IV | 20.1 (2.6) | Omega-3 and placebo (groups were collapsed) | QIDS |
| **Clayton, 2009 (31)** | Uncontrolled open label trial | Australia | 18 | 66.7 | Juvenile bipolar disorder, prescribed with mood-stabilising medication | DSM-IV | F= 16.1 (0.8)  M= 13 (1.1) | Omega-3 (no comparison) | YMRS, HDRS, GASC, CBCL-PR, red blood cell EPA, DPA and DHA levels |
| **Fristad, 2021 (33)** | Observational follow up | United States | 38 | 37.0 | Children with depression participated in OATS study | DSM-IV | 14.6 (2.5) | Those who continued/  initiated omega-3 use following RCT vs. those who did not | CDRS-R, YMRS |
| **McNamara, 2014 (32)** | Open label trial | United States | ~~14~~20 | 60.0 | Adolescents with SSRI-resistant major depressive disorder | DSM-IV | 15.6 (3.2) | High dose vs. low dose omega-3 | CDRS-R, YMRS, erythrocyte fatty acid composition, side effects, blood analyses |

CBCL-PR = Child Behaviour Checklist – Parent Report; CDRS-R = Children’s Depression Rating Scale – Revised; CGIS = Clinical Global Impressions Scale; DHA = docosahexaenoic acid; DPA = Docosapentaenoic acid; DSM-IV = Diagnostic and Statistical Manual of Mental Disorders – Fourth Edition; EPA = eicosapentaenoic acid; F = females; GASC = Global Assessment Scale for Children; HDRS = Hamilton Depression Rating Scale; M = males; OATS = Omega-3 and Therapy study; PRS = parental report of study; QIDS = Quick Inventory of Depressive Symptomatology; SSRI = selective serotonin reuptake inhibitor; YMRS = Young Mania Rating Scale

**Supplementary Table 6. Findings from non-randomised controlled trials investigating the effect of omega-3 supplementation on depression and/or anxiety (n=4)**

| **Study ID** | **Omega-3 daily dose** | **%  EPA, DHA** | **Other interventions**  **(% of participants)** | **Depression** | **Anxiety** | **Other outcomes** | **Side effects and adherence** |
| --- | --- | --- | --- | --- | --- | --- | --- |
| **Amminger, 2015** | 1180mg for 12 weeks | 59.3% EPA  40.7% DHA | Cognitive-behavioural case management (100) | At end of trial, 25% had clinically relevant depressive symptoms. 75% had  improved clinical symptoms (QIDS). | Not studied | Not studied | Not studied |
| **Clayton, 2009** | 1920mg  for 6 weeks | 18.8% EPA  81.2% DHA | Lithium (50),  valproate (39),  quetiapine (11)  Risperidone, dexamphetamine, and SSRIs (unknown) | ↓ depression symptoms (HDRS) | Not studied | Significant improvements in global functioning (GASC). Significant ↓ in mania (YMRS),  internalising and externalising symptoms  (CBCL-PR). Increase in red blood cell EPA and DHA levels and decrease in DPA from pre-post treatment. Changes in mood and behaviour were not significantly correlated with red blood cell LCn-3PUFA levels. | 3 participants withdrew because of gastrointestinal disturbance. Mean adherence = 84.8% ± 4.1, estimated by capsule count-back and interviews among study completers |
| **Fristad, 2021** | Not reported | Not reported | Therapy after RCT (58). Mood stabilisers, anti-obsessionals, antidepressants, anti-psychotics after RCT (63) | ↓ depressive symptoms at follow up for families persisting with omega-3 (CDRS-R) | Not studied | No significant group differences in YMRS scores | Side effects not studied. 50% of follow-up sample reported taking omega-3 after the original RCT. |
| **McNamara, 2014** | 2400mg (low dose) OR   16200mg (high dose) | 66.7% EPA  33.3% DHA | Fluoxetine (50), citalopram (15), escitalopram (5), sertraline (30) | No significant  change in  depression  symptoms in high dose group  compared to low dose group. ↓ depression  symptoms over time for both groups (CDRS-R). | Not studied | Significant decrease in mania symptoms (YMRS) in high dose group. Change in depression symptoms was uncorrelated with change in EPA and DHA levels. | Headache, difficulty staying asleep, diarrhoea, difficulty waking up, dizziness when standing, appetite increase, dizziness, and joint aches greater in low dose group than high dose group. Nausea, lethargy, abdominal pain, vomiting greater in high dose group compared to low dose group. Mean adherence: low dose (92.0%); high dose (97.0%) |

↑ denotes increase in symptoms; ↓ denotes decrease in symptoms; CDRS-R = Children’s Depression Rating Scale – Revised; DHA = docosahexaenoic acid; DLPFC = dorsolateral prefrontal cortex; EPA = eicosapentaenoic acid; GASC = Global Assessment Scale for Children; Glx = glutamine;  Glu = glutamate; LCn-3PUFA = long-chain n-3 polyunsaturated fatty acid; MDD = Major depressive Disorder; QIDS = Quick Inventory of Depressive Symptomatology; RCT = Randomised Controlled Trial; SSRI = selective serotonin reuptake inhibitor; YMRS = Young Mania Rating Scale
